# STOMAPY: Artificial Intelligence for Risk Stratification of Outcomes Requiring Enterostomal Therapy After Hospital Discharge Following Colorectal Surgery

**DOI:** 10.64898/2026.05.11.26352943

**Authors:** Anne Carolline Flores de Souza Barbosa Teixeira, Oneide dos Anjos Pereira, Josielma Prata Vasconcelos, Jordana Maria Freitas Alves, Cláudio Eduardo Corrêa Teixeira

## Abstract

Introduction: Infectious and wound-healing complications after colorectal surgery often increase the complexity of local care and the need for specialized enterostomal therapy follow-up after hospital discharge. Despite the growing use of predictive models in digestive surgery, a translational gap remains between perioperative prediction and the practical organization of specialized care. Therefore, the aim of this study was to develop and temporally validate a machine-learning-based risk stratification model to estimate the probability of post-discharge outcomes associated with greater demand for enterostomal therapy after colorectal surgery. Methods: This was a retrospective observational study including 7,908 patients who underwent colorectal surgery between 2005 and 2014. The outcome was defined as the occurrence of superficial surgical site infection, delayed wound healing, or abdominal sinus formation. Routinely available preoperative and intraoperative variables were used as predictors. The primary model was based on gradient boosting with isotonic calibration. Temporal validation was performed by separating cohorts according to year of surgery. Performance was assessed using ROC-AUC, PR-AUC, Brier score, calibration, and decision-oriented clinical metrics. Clinical utility was examined through percentile-based risk stratification and Decision Curve Analysis (DCA). Results: The outcome prevalence in the test set was 6.6%. The calibrated model achieved a ROC-AUC of 0.64 and a PR-AUC of 0.11, with a Brier score of 0.061. The Top-10% risk stratum concentrated approximately twice the baseline event rate (≈14% vs. 6.6%), with a number needed for intensified follow-up of 7 patients to identify one event. Decision curve analysis showed greater net benefit than strategies of following all or no patients, particularly for threshold probabilities between 3% and 13%. Models based exclusively on preoperative or intraoperative variables performed worse than the combined model. Conclusion: STOMAPY demonstrated the ability to organize patients along a continuous gradient of risk for post-discharge outcomes associated with greater demand for enterostomal therapy. Although discriminatory performance was moderate, the adequate calibration, temporal validation, and net benefit observed across clinically plausible thresholds support its usefulness as a tool for proportional care prioritization rather than as an individual diagnostic test. Prospective studies and external validations are needed to confirm direct clinical impact.

## INTRODUCTION

The creation of intestinal stomas, including ileostomies and colostomies, remains common in different colorectal surgery settings, both as a definitive procedure and as a protective strategy for higher-risk anastomoses and in oncological or inflammatory contexts [1-2]. Although widely used and technically standardized, stoma creation should not be regarded as a simple surgical act, since it is associated with a broad spectrum of early and late complications, with a direct impact on rehabilitation, quality of life, and the use of healthcare resources, especially those related to specialized enterostomal therapy care [3-4].

Stoma-related morbidity is heterogeneous and varies widely across studies, reflecting differences in methodological design, follow-up duration, and outcome definitions. A systematic review found that the incidence of stoma-related complications ranged from 2.9% to 81.1%, with peristomal skin complications and parastomal hernia among the most frequently reported events [4]. These complications are clinically relevant because they often require repeated interventions, including adjustments of ostomy appliances, effluent management, skin and wound care, self-care education, and psychosocial support, thus constituting a central focus of enterostomal therapy practice [3,5].

Among the adverse events associated with stomas, peristomal skin complications are among the most prevalent and costly. Observational evidence and economic analyses indicate that these lesions are common, are associated with a significant increase in costs, and often follow a recurrent course, requiring repeated reassessments and prolonged follow-up [5-6]. Recent international consensus statements have also drawn attention to medical adhesive-related skin injuries in the peristomal area and to the need for standardization in the assessment, prevention, and management of these conditions, reinforcing the role of specialized care in the safe and effective care of people with stomas [7-8]. At the same time, evidence syntheses have identified clinical and therapeutic factors associated with peristomal skin complications after stoma surgery, such as diabetes and other metabolic conditions, but they also highlight methodological limitations and heterogeneity of findings, indicating room for more robust and applicable predictive strategies [8-9].

In this context, it becomes strategically relevant to anticipate, already during the perioperative period, which patients are more likely to develop complications and postoperative trajectories that, in practice, increase the demand for local care and intensified enterostomal therapy support. Landmark studies in colorectal surgery have already explored associations between intraoperative factors and infectious complications occurring within up to 30 days after surgery, such as the registry-based analysis by Walters et al., which investigated the relationship between intraoperative body temperature and postoperative infections [10]. However, an important translational gap remains: transforming routinely collected preoperative and intraoperative variables into risk stratification tools capable of operationally guiding the planning of specialized care, including consultation prioritization, follow-up intensity, targeted therapeutic education, and allocation of specialized nursing time.

Recent literature indicates growing use of machine learning methods to predict postoperative complications in digestive surgery, often with competitive performance compared with traditional statistical models. However, these studies show heterogeneity in design, variable selection, validation strategies, and clinical interpretation, as well as limited integration into care pathways [11-14]. Thus, by proposing an Artificial Intelligence model as an enterostomal therapy-oriented risk stratification approach, based on preoperative and intraoperative variables and on postoperative outcomes that function as proxies for greater enterostomal therapy care complexity in the post-discharge period, this study seeks to help build a bridge between perioperative prediction and the practical organization of specialized enterostomal therapy care.

## METHODS

### Study design and data source

This was a retrospective observational study aimed at the development and temporal validation of a multivariable prediction model for the individual estimation of the probability of outcomes associated with greater demand for enterostomal therapy after hospital discharge following colorectal surgery. The methodological conduct was structured in accordance with the TRIPOD principles for clinical prediction models [13,15].

The data used in this study were obtained from the Colorectal Surgery Database and the Cleveland Clinic Perioperative Health Documentation System, and are available under a CC BY-NC-SA license at https://www.causeweb.org/tshs/core-temperature/, as well as at https://www.kaggle.com/datasets/mexwell/core-temperature-during-surgery/data. These datasets include variables from 7,908 adult patients who underwent colorectal surgery between 2005 and 2014, with sufficient in-hospital and post-discharge follow-up for the assessment of postoperative outcomes.

### Study population

All patients underwent colorectal surgery under general anesthesia, with a duration longer than 60 minutes and continuous monitoring of core body temperature via an esophageal probe. No additional exclusion criteria were applied beyond the unavailability of essential information, in order to preserve clinical representativeness and minimize selection bias.

### Outcome definition

The outcome was defined as a binary variable representing the occurrence of at least one event related to surgical wound integrity and the healing process, including delayed healing, superficial surgical site infection, surgical wound infection, and abdominal sinus formation.

Although these components are etiologically heterogeneous, they were grouped because they share a common care-related convergence: all imply increased complexity of local care and a higher likelihood of requiring specialized enterostomal therapy follow-up in the post-discharge period. The outcome was operationalized as 1 in the presence of any event and 0 in the absence of events.

### Predictor variables

Candidate predictors were defined *a priori* based on clinical plausibility and availability within real-world care pathways. Preoperative variables included age, sex, body mass index, Charlson score, congestive heart failure, diabetes, renal failure, liver disease, neoplasia, coagulopathies, fluid and electrolyte disorders, weight loss, anemia, prior steroid use, immunosuppressive drug use, and history of drug abuse. Intraoperative variables included duration of surgery, type of surgical approach, and body temperature measures, including time-weighted mean temperature and temperature at the end of the procedure.

No prior univariate selection was performed, in order to avoid filtering bias based on marginal associations and to preserve coherence between the exploratory and confirmatory stages. In addition, risk of bias was assessed in light of the recommendations of PROBAST [16].

### Data preparation and handling of missing data

Missing values in numerical variables were handled by simple median imputation, implemented exclusively within the training pipeline. This strategy was considered appropriate given the low proportion of missing data and the need for compatibility with both linear models and tree-based models. In addition, binary variables were already originally coded as 0/1 in the available dataset.

### Data splitting strategy and temporal validation

Validation followed a strict temporal strategy. Patients operated on in earlier years comprised the development set, whereas patients from subsequent years were reserved exclusively for testing. The test set remained completely isolated throughout all stages of development, including hyperparameter tuning and calibration. This approach simulates a prospective clinical implementation scenario and reduces the risk of optimistic overestimation of performance.

### Statistical modeling

The modeling was formalized as the estimation of the conditional probability P(Y = 1 | X), where Y represents the composite outcome and X the predictor vector. Penalized logistic regression with Elastic Net was initially used as the reference model, imposing L1 and L2 regularization to control multicollinearity and mitigate overfitting. The main model was based on decision trees optimized by gradient boosting (HistGradientBoostingClassifier), chosen for its ability to capture nonlinear relationships and implicit interactions among heterogeneous clinical variables. Hyperparameter optimization was conducted by year-grouped cross-validation within the development set, restricting functional complexity through limits on tree depth, number of terminal nodes, and learning rate, with the aim of promoting stability and generalization.

### Probability calibration

Given the probabilistic nature of the clinical application, isotonic regression calibration was performed on the development set. Probabilistic adequacy was assessed using the Brier score and the slope and intercept parameters obtained by logistic regression between the logit of the predicted probability and the observed outcome.

### Performance assessment

Model performance was evaluated exclusively on the temporal test set. Discrimination was quantified by the area under the ROC curve and the area under the precision–recall curve. Calibration was examined using the Brier score and calibration regression parameters. In addition, decision-oriented metrics were calculated under operational policies based on upper percentiles of the risk distribution (Top-k), including sensitivity, specificity, predictive values, lift, and number needed for focused clinical follow-up. The model’s clinical utility was evaluated through decision curve analysis, estimating net benefit across different threshold probabilities.

### Operational risk stratification

The estimated continuous probability was used to construct risk strata based on quantiles derived exclusively from the development set. This approach avoids arbitrary fixed thresholds and allows prioritization to be aligned with the operational capacity of the enterostomal therapy service. The model was designed as a population-ranking and proportional resource-allocation tool, rather than as an individual binary diagnostic test.

### Robustness, reproducibility, and overfitting control

Explicit strategies were adopted to mitigate overfitting, including penalized regularization, restriction of the complexity of the gradient-based model, temporal validation, and calibration restricted to the development set. The pipeline was structured deterministically, with fixed random seeds and full serialization to ensure computational reproducibility.

### Methodological limitations

Despite temporal validation, the study remains limited to a single center, which may restrict external generalizability. Simple imputation does not incorporate the uncertainty associated with multiple imputation. Therefore, external multicenter validation represents a necessary future step.

## RESULTS

In the independent temporal test set, the STOMAPY model showed consistent performance, with moderate discrimination, adequate calibration, and measurable operational utility, concentrating events in the higher-risk strata, generating net clinical benefit at thresholds compatible with remote monitoring, and functioning as a population-ranking tool. In other words, the model proved useful as a tool to support rational prioritization in enterostomal therapy after hospital discharge following colorectal surgery.

### General characteristics of the study population

A total of 7,908 patients who underwent colorectal surgery were analyzed. Considering the composite outcome restricted to events more frequently observed in the post-discharge period (superficial surgical site infection, wound infection, delayed healing, or abdominal sinus), the overall prevalence was 7.6% (598/7,908).

In the temporal split, 5,085 patients comprised the development set and 2,823 the test set. In the test set, the prevalence was approximately 6.6%. Patients with outcome occurrence had a higher body mass index (28.9 ± 6.9 vs. 26.4 ± 6.1), longer surgical duration (median 238 vs. 191 minutes), and a slightly higher comorbidity burden (median Charlson score 1 vs. 0). The proportion of open surgery was similar between groups. Mean intraoperative temperature showed only a small absolute difference, without clinically relevant magnitude. These findings reinforce that post-discharge risk associated with greater demand for enterostomal therapy is multifactorial and is not determined by a single isolated variable.

### Discriminative performance of the STOMAPY model

After parsimony-based reduction and isotonic calibration, the STOMAPY model achieved an ROC-AUC of 0.64 (Figure 1), indicating moderate discriminative ability; that is, the model is able, on average, to assign a higher probability of the event to patients who will in fact experience the outcome compared with those who will not.

**Figure 1.**
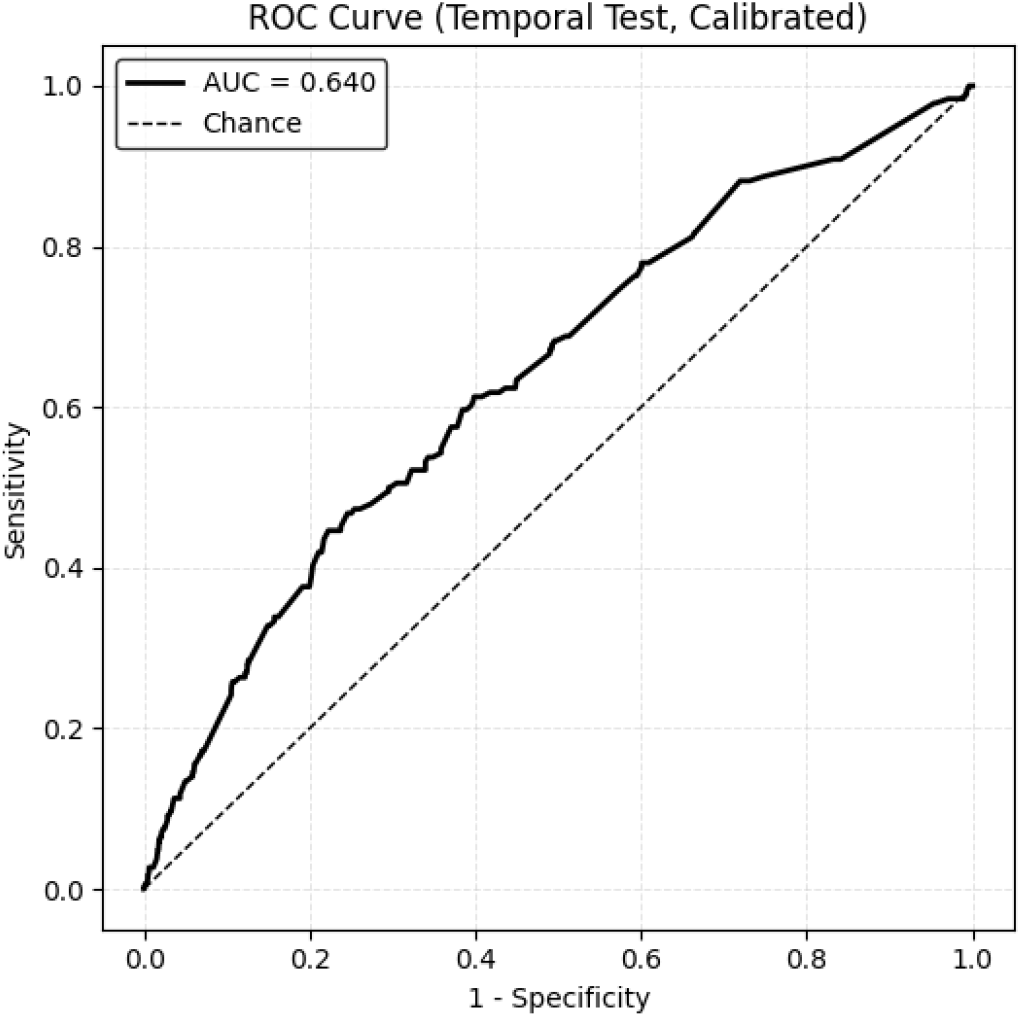
ROC curve of the STOMAPY model in the temporal test set (calibrated probabilities). The model achieved an AUC of 0.640, indicating moderate discriminative ability to distinguish patients who developed post-discharge outcomes associated with greater demand for enterostomal therapy from those who did not experience such events.

The PR-AUC is particularly relevant in low-prevalence scenarios (as in the present study, with approximately 6–8%), because it evaluates precision in identifying positive events. The value obtained (0.11) substantially exceeds the baseline prevalence, indicating informational gain. Likewise, the mean squared error between predicted probabilities and observed outcomes, or Brier score, remained low (0.061), indicating good overall probabilistic accuracy.

**Table 1.** Clinical and surgical characteristics of patients undergoing colorectal surgery, according to the occurrence of the proxy outcome associated with greater demand for intensified enterostomal therapy care. Cleveland Clinic, 2005–2014.

| Variable | Total<br>(n = 7.908) | W/out proxy outcome<br>(n [%] = 7.310 [92,4]) | W/ proxy outcome<br>(n [%] = 598 [7,6]) |
| --- | --- | --- | --- |
| Age, yrs. - average (s.d.) | 55.7 (16.8) | 55.7 (16.9) | 56.8 (15.5) |
| Female - n (%) | 4,002 (50.6) | 3,688 (50.6) | 314 (51.0) |
| BMI - average (s.d.) | 26.6 (6.2) | 26.4 (6.1) | 28.9 (6.9) |
| Charlson Comorbidity Index -<br>median (IQR) | 1 (0-2) | 0 (0-2) | 1 (0-2) |
| Surgery duration, minutes -<br>median (IQR) | 195<br>(135-269) | 191<br>(134-265) | 238<br>(175-311) |
| Open surgery - n (%) | 6,504 (82.2) | 6,006 (82.1) | 498 (83.3) |
| Mean intraoperative<br>temperature (°C) –<br>average (s.d.) | 36.02 (0.53) | 36.02 (0.53) | 36.1 (0.54) |

s.d., standard deviation; IQR = interquartile range. The proxy outcome represents the occurrence of at least one of the following events: superficial surgical site infection, wound infection, fascial dehiscence, delayed healing, or abdominal sinus formation.

In practical terms, these measures show that the model organizes patients along a risk scale. Patients who actually developed complications tended, on average, to receive higher risk scores than those who did not. An ROC-AUC value of 0.64 indicates that this ranking performs better than chance and is clinically useful for prioritization purposes.

### Probabilistic calibration

The calibration curve demonstrated adequate agreement between predicted risk and observed risk across clinically relevant ranges (Figure 2). In other words, when the model estimates, for example, a risk of approximately 10%, the actual observed proportion of events tends to be close to that value. For healthcare professionals, this means that the probabilities provided by the model can be interpreted directly, without a relevant systematic tendency toward overestimation or underestimation.

**Figure 2.**
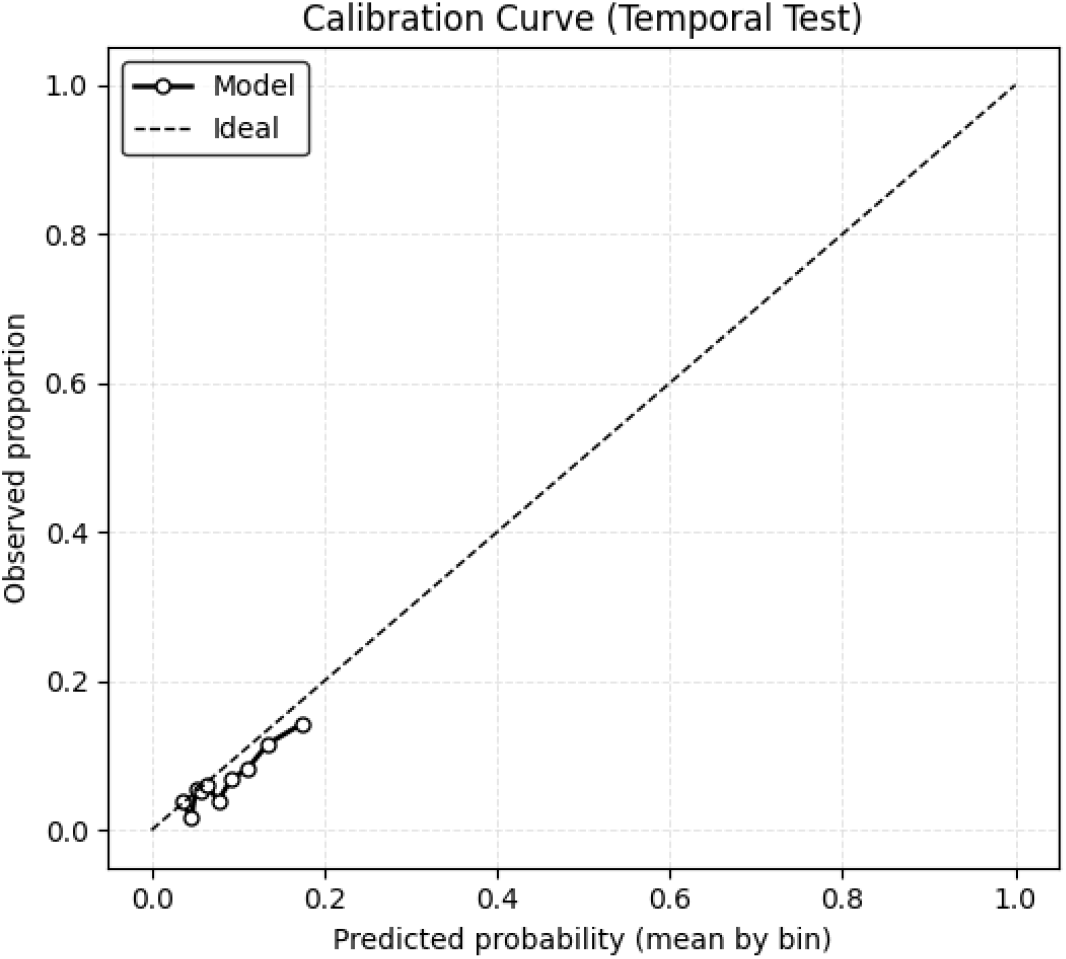
Calibration curve of the STOMAPY model in the temporal test set. The figure shows the relationship between the mean probability predicted by the model (horizontal axis) and the observed proportion of events in each risk stratum (vertical axis), calculated by grouping predictions into probability quantiles. The diagonal dashed line represents perfect calibration (predicted probability equal to observed probability). The points demonstrate good agreement across clinically relevant risk ranges (approximately 3% to 20%).

### Comparison between preoperative and pre + intraoperative models

The model based exclusively on preoperative data showed an ROC-AUC close to 0.59. The model based exclusively on intraoperative data showed similar performance (approximately 0.60). The integration of preoperative and intraoperative data increased the predictive capacity of the model, with the ROC-AUC rising to approximately 0.64. This incremental gain demonstrates that combining prior clinical information with intraoperative data improves risk discrimination, although neither of these dimensions alone is sufficiently strong to achieve high accuracy. Thus, the result of this analysis indicates that the risk of post-discharge complications depends not only on the patient’s clinical profile, nor only on what occurs during surgery, but on the combination of both.

### Operational risk stratification

Because the aim of STOMAPY is to support care organization, the results were analyzed under prioritization policies based on follow-up capacity. When prioritizing the 5% of patients with the highest predicted risk, 12% of all events were captured by the model, with an event rate of 15.5% in the prioritized group, whereas the event rate in the remaining patients was 6.1%. When capacity was expanded to the 10%, 20%, and 30% of patients with the highest predicted risk, the model captured 20.9%, 37.6%, and 48.4% of all events, respectively, with event rates in the prioritized group of 13.8%, 12.4%, and 10.6%, while the event rates in the remaining patients were 5.8%, 5.1%, and 4.9%, respectively (Figure 3, upper panels).

**Figure 3.**
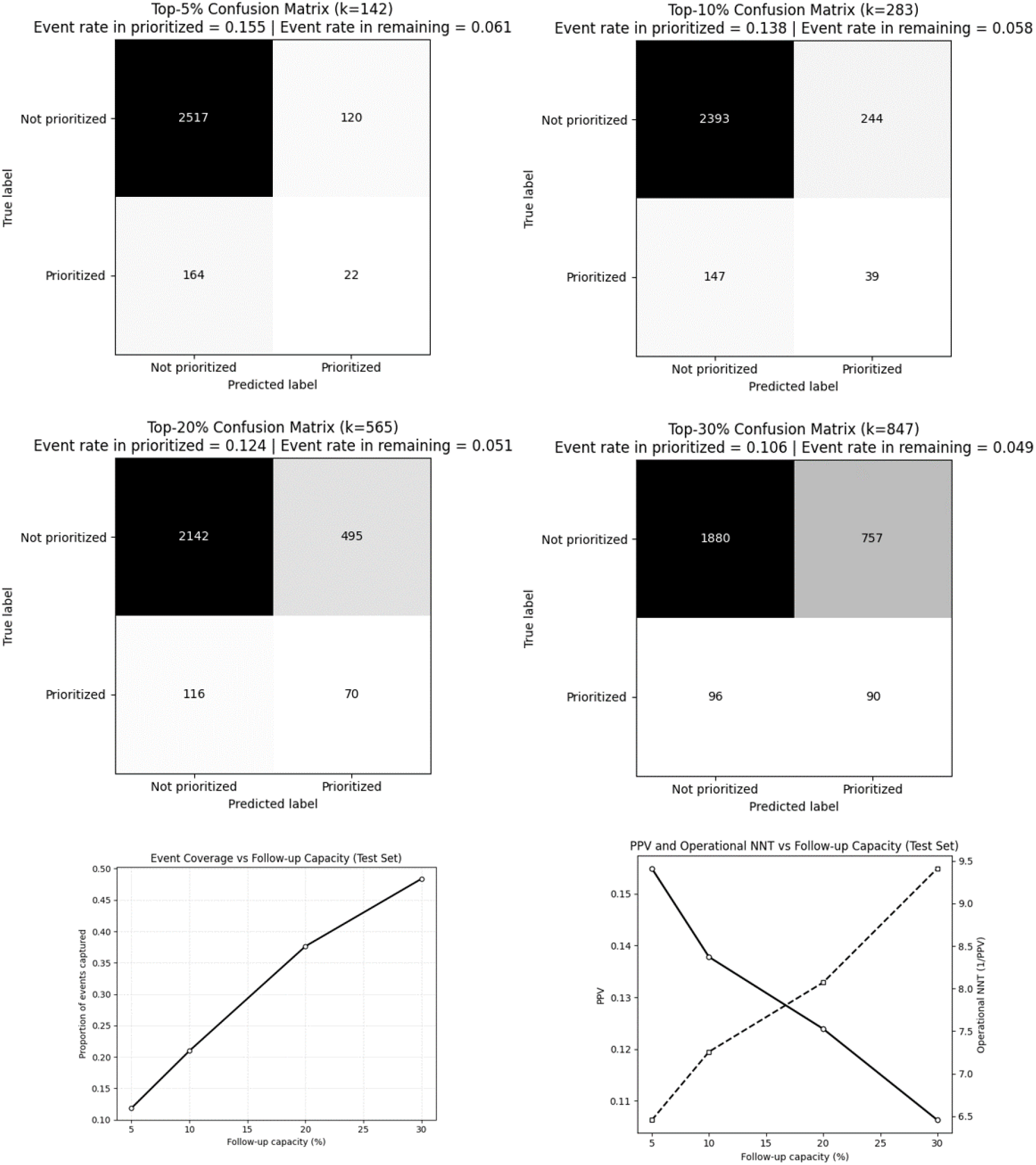
Operational risk stratification by the STOMAPY model in the temporal test set. The four upper panels present the confusion matrices corresponding to prioritization strategies based on the upper percentiles of the predicted risk distribution (Top-5%, Top-10%, Top-20%, and Top-30%). In each scenario, patients classified as “Prioritize” correspond to those with the highest estimated risk, according to the simulated care capacity. The lower panels show the proportion of events captured as a function of follow-up capacity (left) and the relationship between positive predictive value (PPV) and the number needed for intensified follow-up (operational NNT) according to follow-up capacity.

On the other hand, the number needed for prioritized follow-up (operational NNT) was approximately 7, meaning that by prioritizing follow-up for 7 patients predicted to be at highest risk, 1 event is identified on average. A progressive reduction in positive predictive value was also observed as coverage increased, which is an expected phenomenon when patients with progressively lower risk are included (Figure 3, lower panel).

Overall, these results indicate that the model functions as a tool that ranks the risk of post-discharge outcomes requiring enterostomal therapy from “most likely” to “least likely.” If the service has the capacity to follow only 10% of patients after hospital discharge, the model will concentrate approximately twice the event rate in this group compared with the general population. If the service can follow 30% of patients after hospital discharge, nearly half of all events will be included, but with lower proportional concentration.

### Performance at a fixed 10% threshold

Considering a fixed probability threshold of 10%, Figure 4 shows that the model was able to predict 89 true positives (patients correctly identified as high risk who developed the outcome), 1,918 true negatives, 97 false negatives, and 719 false positives. At this threshold, the model achieved an approximate sensitivity of 48%, specificity of 73%, positive predictive value of about 11%, and negative predictive value close to 95%.

**Figure 4.**
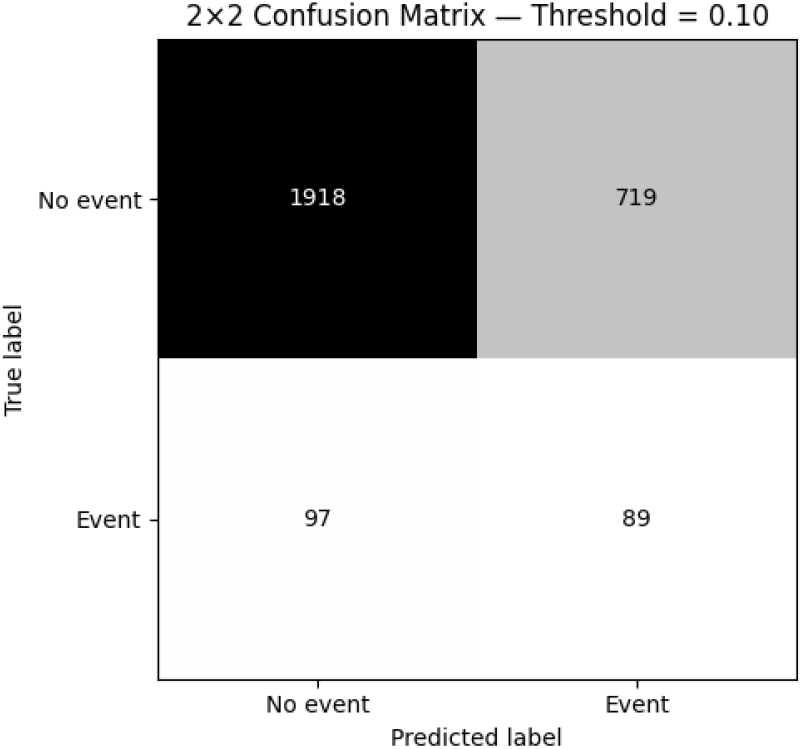
Confusion matrix of the STOMAPY model in the temporal test set (probability threshold = 10%). The 2×2 matrix presents the classification of patients considering a predicted risk ≥10% as an indication for prioritization for focused enterostomal therapy follow-up.

These results reinforce that the model is particularly effective in identifying low-risk patients (high NPV), being able to correctly identify approximately half of the patients who will develop the outcome, and therefore being more suitable for population ranking and stratification than for rigid individual binary classification. In other words, the model appears to be more effective at identifying low-risk patients than at confirming high risk.

### Decision Curve Analysis

The model shows greater net benefit than the strategies of following all patients and following none, particularly within the threshold range of approximately 3% to 13% (Figure 5, upper panel). For higher thresholds (>15%), the net benefit approaches zero, suggesting that the model is more suitable for proportional prioritization strategies than for rigid binary decisions based on high thresholds.

**Figure 5.**
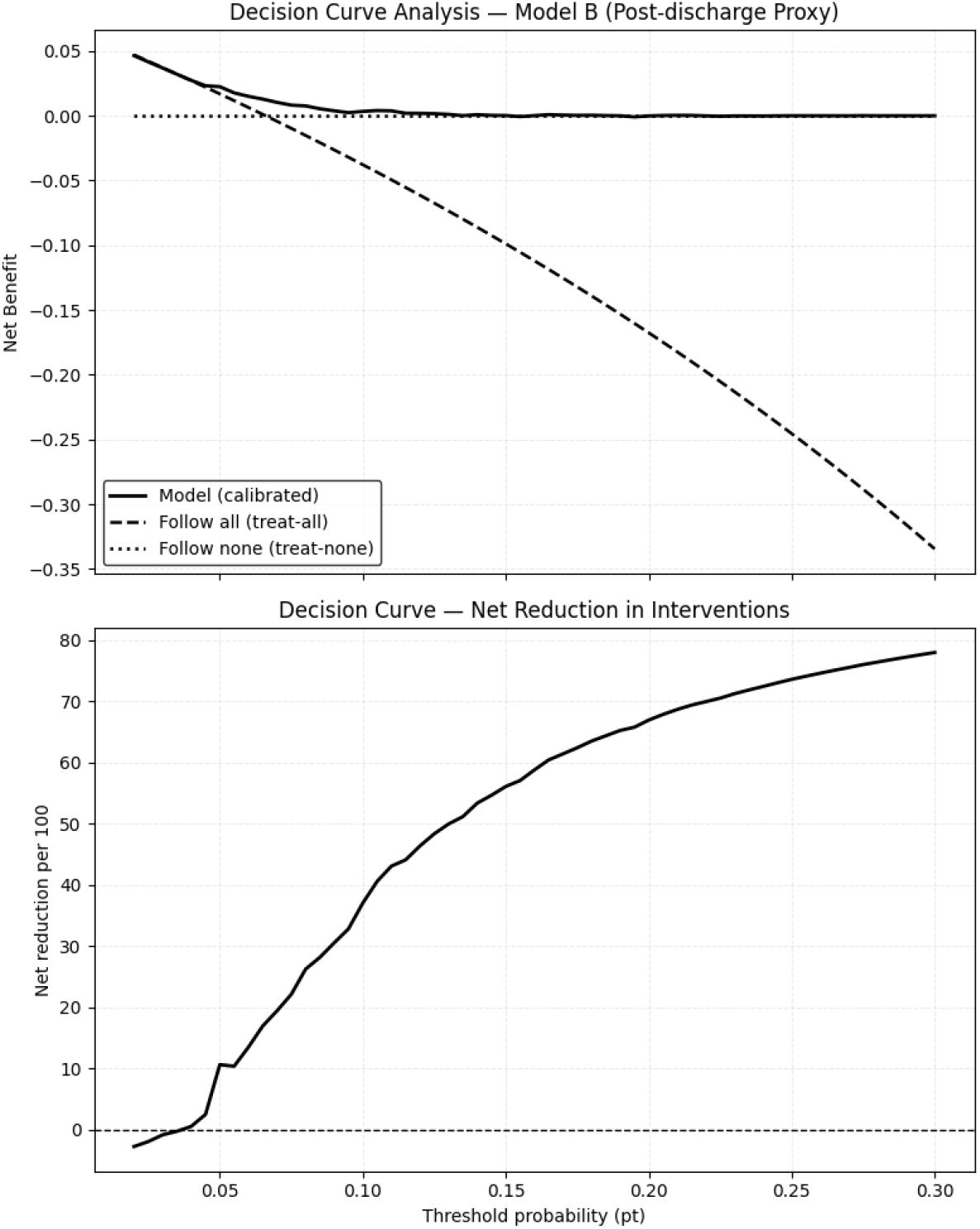
Decision Curve Analysis of the STOMAPY model in the temporal test set. Upper panel: Net benefit comparing three strategies: use of the calibrated model (blue line), universal follow-up (treat-all, dashed orange line), and no systematic follow-up (treat-none, dotted green line). Lower panel: Net reduction in unnecessary interventions per 100 patients, according to the threshold probability (pt).

In addition, for thresholds between approximately 3% and 13%, use of the model allows an increasing number of unnecessary focused follow-ups to be avoided, without a proportional loss in event identification (Figure 5, lower panel). As the threshold increases, the net reduction exceeds 70 interventions per 100 patients at higher thresholds, reflecting greater operational specificity. In other words, at higher thresholds, the model makes it possible to avoid a substantial number of unnecessary follow-ups without proportionally compromising event detection.

Thus, the results indicate that the STOMAPY model appears to be most useful when applied to low-risk, low-cost intervention strategies, such as structured post-discharge telemonitoring, reinforced education, or scheduled telephone contact. STOMAPY should be interpreted as a tool for proportional prioritization of healthcare resources, rather than as an individual binary decision-making instrument. Furthermore, because the model’s performance was evaluated in a temporally distinct cohort, these results reinforce the model’s stability in the face of variations in clinical practice. Therefore, the model also proved capable of generating measurable clinical gain when used to organize post-discharge enterostomal therapy follow-up in a stratified manner aligned with the available care capacity.

## DISCUSSION

The present study proposes and validates a machine-learning-based risk stratification strategy to estimate the probability of postoperative trajectories associated with greater demand for local care, wound management, and specialized follow-up after colorectal surgery, in clinical contexts in which colostomy is frequently used. The main contribution of this work does not lie in the isolated maximization of algorithmic performance, but rather in the delivery of continuous, calibrated, and clinically actionable risk stratification, with the potential to support the organization of enterostomal therapy care and care prioritization in settings with limited specialized resources.

The discriminative performance of the model used was moderate (AUC ≈ 0.64), a value compatible with the multifactorial and partially stochastic nature of postoperative complications. In surgery, however, the clinical usefulness of a model should not be judged exclusively by the magnitude of the AUC, but also by its calibration, decision impact, and ability to support care organization [17-18]. Nevertheless, the model was able to concentrate approximately twice the event rate in the upper risk stratum (Top-10%) compared with the baseline prevalence, demonstrating practical utility in scenarios of proportional prioritization.

Decision curve analysis showed greater net benefit than the extreme strategies of following all patients or none after hospital discharge, especially for thresholds between 3% and 13%, a range compatible with low-risk monitoring strategies such as telemonitoring and structured post-discharge follow-up. Decision curve analysis makes it possible to assess the net clinical benefit of predictive models by integrating sensitivity, specificity, and implicit risk preferences across different decision thresholds [19-21].

### Interpretation of the main findings

The comparison between models based exclusively on preoperative information and those incorporating intraoperative variables showed a consistent performance gain with the inclusion of the latter, particularly in terms of discrimination and precision for low-prevalence events. This finding is consistent with the literature showing that postoperative complications in colorectal surgery reflect both the patient’s baseline profile and indirect markers of surgical complexity and stress throughout the procedure. Previous studies also indicate that procedure duration and intraoperative physiological conditions, such as thermal changes, are associated with a greater risk of infectious and wound-healing complications [10,22-23].

From a care perspective, this feature allows the tool to be used at two complementary time points: preoperatively, for initial resource planning, patient education, and follow-up organization; and in the immediate postoperative period, after incorporation of intraoperative data, for dynamic risk updating and adjustment of the intensity of enterostomal therapy follow-up. This logic is compatible with contemporary models of personalized perioperative care and with current recommendations for integrating predictive models into real clinical practice [13-15].

### Role of predictors and clinical plausibility

The analysis of the relative importance of variables in the final model indicated that factors related to surgical complexity and metabolic vulnerability contributed most to risk stratification. Among the predictors, surgical duration, body mass index, and age stood out, followed by intraoperative markers such as mean intraoperative temperature and type of surgical approach. Variables associated with fluid and electrolyte disorders and systemic comorbidities contributed additionally, although with less relative weight.

This hierarchy is clinically plausible, since longer procedures and patients with greater metabolic frailty tend to have a higher risk of infectious and wound-healing complications, scenarios in which the demand for frequent dressings, skin surveillance, and specialized reassessments is usually intensified [10,22-23]. It is important to emphasize that variable importance was used exclusively for parsimony and model stability purposes and should not be interpreted as a measure of individual causal effect, in accordance with good practices for the development and interpretation of clinical predictive models [14-15].

### Relevance to enterostomal therapy and use of a proxy outcome

Although the database analyzed does not contain enterostomal therapy-specific variables, there is clinical plausibility in using infectious and wound-healing complications as proxy outcomes for contexts of greater demand for enterostomal therapy care. Superficial infectious complications, delayed healing, and abdominal sinus formation represent clinical contexts in which the complexity of local care and the demand for specialized enterostomal therapy follow-up tend to be substantially increased [2,8,24].

Observational studies and systematic reviews show that complications related to the stoma and peristomal skin remain frequent, costly, and associated with poorer quality of life, as well as greater use of healthcare services [3,5-6,9]. Thus, although the adopted outcome does not directly represent classic enterostomal therapy events, it captures a clinical context of greater care complexity, in which the enterostomal therapist’s role tends to be more intensive, prolonged, and central to patient rehabilitation.

### Clinical utility of risk stratification

A central finding of the study was the model’s ability to concentrate events in smaller subgroups, making it possible to express the efficiency of stratification through the number of patients who need to be prioritized in order to identify one event of the composite outcome. This metric does not imply direct therapeutic benefit, but pragmatically translates the organizational utility of the model. Unlike AUC, which assesses statistical discrimination, net benefit considers the relationship between true positives and false positives weighted by the clinical threshold adopted, thereby approximating real care decisions [19-20].

The literature shows that stoma-related complications, including readmissions for dehydration, difficulties in effluent management, and wound problems, are frequent causes of unplanned service use after colorectal surgery [25-27]. These findings reinforce the importance of targeted follow-up strategies and support the potential of risk stratification as a tool to aid prioritization of enterostomal therapy care, without replacing individual clinical assessment.

### Methodological aspects and alignment with current guidelines

From a methodological standpoint, the study is aligned with current recommendations for clinical predictive models by emphasizing temporal validation, explicit calibration assessment, and the use of appropriate metrics for low-prevalence outcomes, such as the area under the precision-recall curve and the Brier score [13-15,28]. The decision not to prioritize the ROC curve exclusively reflects a concern with real clinical utility, recognizing that discrimination alone may overestimate practical relevance when not contextualized in terms of net benefit and decision thresholds [17,19-21]. Risk of bias assessment was conducted in light of PROBAST recommendations, reinforcing the methodological transparency of the study [16]. In addition, the use of decision curve analysis in surgical contexts has been recommended as a complementary tool to traditional performance assessment, particularly when the objective of the model is to support organizational decisions or care prioritization [20,29]. Finally, evaluation in a temporally distinct cohort reinforces the model’s stability in the face of changes in surgical practice over the years, conceptually approximating institutional external validation.

### Limitations and recommendations

This study has important limitations. First, the use of a proxy outcome may introduce misclassification error, since it does not directly measure specific enterostomal therapy complications, such as peristomal dermatitis, retraction, or parastomal hernia [2,24]. Second, the absence of detailed information on stoma type, preoperative stoma site marking, devices used, and educational interventions limits more specific analyses for enterostomal therapy practice.

In addition, the data derive from a single institutional system, which may restrict the external generalizability of the findings. Temporal changes in surgical and care practices may also generate data drift, requiring periodic reevaluation of the model. Finally, discriminative performance was moderate, which is expected given the multifactorial nature of the outcome and reinforces that the tool should be used as support for prioritization, rather than as a deterministic criterion.

Based on the findings, external validation of the model in other cohorts is recommended, with eventual local recalibration [15,28]. Future studies should incorporate variables directly related to enterostomal therapy, such as stoma type, site characteristics, and follow-up strategies, as well as prospectively evaluate the impact of stratification on care organization, resource use, and the occurrence of complications [9,11,13-14].

## CONCLUSION

This study demonstrates that a machine-learning-based risk stratification strategy, using routinely available preoperative and intraoperative clinical variables, is capable of organizing patients undergoing colorectal surgery along a continuous gradient of risk for post-discharge outcomes associated with greater local care complexity and potentially increased demand for enterostomal therapy.

The moderate discriminative performance, together with adequate calibration and independent temporal validation, supports its applicability as a tool for proportional care prioritization rather than as an individual binary diagnostic test.

By concentrating events in higher-risk subsets and demonstrating net benefit at thresholds compatible with structured monitoring strategies, STOMAPY provides a solid methodological basis for more efficient organization of post-discharge care in colorectal surgery. Prospective studies and external validations are needed to confirm direct clinical impact and incorporate stoma-specific variables.

## Data Availability

All data produced are available online at https://www.causeweb.org/tshs/core-temperature/

## Notes

### Competing Interest Statement

The authors have declared no competing interest.

### Funding Statement

This study did not receive any funding.

### Author Declarations

The study used (or will use) ONLY openly available human data that were originally located at: https://www.causeweb.org/tshs/core-temperature/

